# Mitochondrial heteroplasmy improves risk prediction for myeloid neoplasms

**DOI:** 10.1101/2024.04.07.24305454

**Authors:** Yun Soo Hong, Sergiu Pasca, Wen Shi, Daniela Puiu, Nicole J Lake, Monkol Lek, Meng Ru, Megan Grove, Anna Prizment, Corinne E. Joshu, Elizabeth A. Platz, Eliseo Guallar, Dan E. Arking, Lukasz P. Gondek

**Author notes:** These authors contributed equally (Yun Soo Hong, Sergiu Pasca). These authors contributed equally as co-corresponding authors (Dan E. Arking, Lukasz P. Gondek). Competing Interests: The authors declare no competing financial interests. **Correspondence:** Lukasz P. Gondek, Johns Hopkins University School of Medicine 1650 Orleans St, CRB1-290, Baltimore, MD 21287, Dan E. Arking, Johns Hopkins University School of Medicine 733 N. Broadway Street, MRB-459 Baltimore, MD 21205.

## Abstract

The most well-known pathogenic risk factor for myeloid neoplasms (MN) is clonal hematopoiesis of indeterminate potential (CHIP) ^1^. However, MN can develop in CHIP negative individuals, indicating that additional markers of clonal expansion might also be informative. Heteroplasmy, defined as the presence of mitochondrial DNA (mtDNA) mutations in a subset of cellular mtDNA, has been associated with hematological malignancies ^2^ and could represent a marker of clonal expansion ^3^. However, the relationship between heteroplamsy and CHIP, as well as its association with the incidence of MN in the general population is not known. In this study, we explored the association between somatic mtDNA and nuclear DNA (nDNA) mutations (mito-nuclear interaction), its impact on MN incidence, and whether its inclusion to the latest CHIP-based MN prediction algorithm could improve risk stratification in over 440,000 participants in the UK Biobank and Atherosclerosis Risk in Communities (ARIC) studies. We found that heteroplasmy count and heteroplasmic variants predicted to be more deleterious were enriched in individuals with CHIP, particularly in those with significantly expanded clones (VAF ≥20%), with more than one CHIP mutation, and with mutations in the spliceosome machinery. Individuals with both heteroplasmy and CHIP were more likely to develop MN than participants with either entity alone. Furthermore, we found a significant and independent association of predicted pathogenic effect of heteroplasmic variants with incident MN, suggesting a causal role of mtDNA variations in MN pathogenesis, even in the absence of CHIP. Finally, incorporating heteroplasmy into an existing risk score model for MN in individuals with CHIP significantly improved the sensitivity by 13.1% and identified 34.4% more cases in the high-risk group (10-year risk ≥10%). In sum, our findings suggest that heteroplasmy, in addition to being a marker of clonal expansion, may be a causal biomarker of MN development, with clinical utility in the general population.

## Introduction

Screening, detection, and control of early malignancy or premalignant lesions are the hallmark of secondary prevention of cancer. This set of interventions, while successfully applied in various solid tumor malignancies, has not been widely adopted in myeloid neoplasm (MN), likely due to the lack of precise biomarkers of MN risk ^4–6^. In recent years, significant progress has been made, fueled mainly by advances in cancer genomics and next-generation sequencing (NGS). The broad application of this technology to large population-based cohorts, providing easy access to sequencing data for the scientific community, allowed for the identification of clonal expansion of hematopoietic cells, also known as clonal hematopoiesis (CH), in over 10% of the adult population^1,7–9^. The most studied form of CH is clonal hematopoiesis of indeterminant potential (CHIP), which is defined as the presence of cancer-associated somatic mutations in hematopoietic cells in otherwise healthy individuals. Given its clonal nature, it is not surprising that some forms of CHIP are myeloid premalignant conditions. Most recently, novel multiparameter MN prediction models including somatic mutations, hematologic indices, and demographic data have been developed to identify the population with CHIP at high risk for developing MN ^10,11^. Unfortunately, these risk scoring systems are limited to individuals with CHIP, who constitute only a small fraction of individuals at risk.

Somatic mitochondrial DNA (mtDNA) mutations are common and have been reported not only in mitochondrial diseases, but also in aging and cancer ^12,13^. Unlike diploid nuclear DNA, mtDNA exists in 10s to 1000s of copies within each cell. Thus, mutations in the mtDNA can exist in a subset of the total cellular mtDNA, a condition termed “mitochondrial heteroplasmy”. As mtDNA repair machinery is limited and not as efficient as the nuclear DNA (nDNA) repair system, the mtDNA mutation rate is 10-17 fold higher than in nDNA ^14,15^. Heteroplasmy can therefore be used as an endogenous cell barcode allowing for lineage tracing and assessment of the clonal expansion of hematopoietic cells and serves as an excellent marker of CH ^3^. In participants from the UK Biobank (UKB), heteroplasmy was present in 30% of individuals, and consistent with CH, the frequency increased with age ^2^. Given their central role in essential cellular processes, mitochondrial alterations are key components of several hallmarks of cancer such as cellular energetics, proliferation, and apoptosis ^16,17^. Thus, it is not surprising that heteroplasmy may provide additional mechanisms of selective growth advantage of abnormal clone(s) and further shape the genomic landscape of CH and MN. We have recently reported that mtDNA heteroplasmy was associated with a 1.5-fold increase in all-cause mortality, and the presence of mtDNA mutations at highly constrained sites was associated with a 4-fold increase in mortality due to leukemia ^2^.

In the current study we used two large population-based cohorts, UKB and Atherosclerosis Risk in Communities (ARIC) study, to examine the association between somatic mtDNA and nDNA mutations (mito-nuclear interaction) in clonal evolution and its impact on the incidence of MN. We further assessed the role of heteroplasmy as a novel predictor of MN risk and whether the inclusion of heteroplasmy into the latest MN prediction algorithms could improve risk classification.

## Results

### Demographics and molecular characteristics of the UKB and ARIC cohorts

The study included a total of 441,936 participants from the UKB (N = 434,304) and ARIC (N = 7,632) cohorts, who had information on both CHIP and mitochondrial heteroplasmy. In the UKB, the mean age was 56.5 (8.1) years, 45.8% (n = 199,046) were men and 94.8% (n = 410,313) were self-identified as White. In ARIC, the mean age was 57.9 (6.0) years, 45.2% (n = 3,453) were men and 76.2% (n = 5,817) and 23.8% (n = 1,815) were self-identified as Whites and Blacks, respectively (Table 1).

**Table 1.** Participant characteristics by presence of CHIP in each cohort.

|  | UK Biobank |  |  | ARIC |  |  |
| --- | --- | --- | --- | --- | --- | --- |
|  | CHIP (-) | CHIP (+) | *P* | CHIP (-) | CHIP (+) | *P* |
| **Number** | 400,707 | 33,597 |  | 7,257 | 375 |  |
| **Age, years** | 56.3 (8.1) | 58.9 (7.6) | <0.0001 | 57.8 (6.0) | 59.6 (6.1) | <0.0001 |
| **Men, %** | 45.9 | 44.8 | <0.0001 | 45.1 | 48.8 | 0.17 |
| **Race, %** |  |  | <0.0001 |  |  | <0.01 |
| **White** | 94.7 | 95.8 | <0.0001 | 76.5 | 70.1 | <0.01 |
| **Black** | 1.6 | 1.3 |  | 23.5 | 29.9 |  |
| **Asian** | 2.2 | 1.8 |  | 0 | 0 |  |
| **Other** | 1.5 | 1.2 |  | 0 | 0 |  |
| **Ever smoker, %** | 59.5 | 62.4 | <0.0001 | 60.5 | 61.6 | 0.70 |
| **Anemia, %** | 4.2 | 4.5 | <0.01 | 9.3 | 12.5 | 0.06 |
| **Thrombocytopenia, %** | 2.2 | 2.5 | <0.001 | 2.1 | 4.9 | 0.01 |
| **Neutropenia, %** | 1.0 | 1.2 | 0.01 | 3.1 | 1.9 | 0.63 |
| **Cytopenia, %** | 7.0 | 7.6 | <0.0001 | 11.1 | 13.7 | 0.30 |
| **MCV, fL** | 91.1 (4.6) | 91.3 (4.6) | <0.0001 | 89.8 (5.1) | 89.6 (5.5) | 0.54 |
| **RDW, %** | 13.5 (1.0) | 13.6 (1.1) | <0.0001 | NA | NA | NA |
| **History of cancer, %** | 6.9 | 8.8 | <0.0001 | 0 | 0 | NA |
| **Heteroplasmy, %** | 27.8 | 34.1 | <0.0001 | 27.9 | 35.2 | <0.01 |

Continuous variables are displayed as either mean (SD) or median (25th and 75th percentiles). Participant characteristics for both cohorts were based on the time of DNA collection. In ARIC, only participants without a cancer diagnosis at the time of blood collection were included in the analysis. Because of missing blood parameters, in ARIC, we were able to evaluate anemia in 6,473 (84.8%) participants; thrombocytopenia in 5,161 (67.6%) participants; neutropenia in 3,372 (44.2%) participants and cytopenia in 3,344 (43.8%) participants. Abbreviations: ARIC, Atherosclerosis Risk in Communities study; CHIP, clonal hematopoiesis of indeterminate potential; MCV, mean corpuscular volume; mMSS, mitochondrial local constraint (MLC) score sum; NA, not available; and RDW, red cell distribution width.

Using whole exome sequencing data (WES), we identified 37,089 CHIP mutations in the UKB and 435 CHIP mutations in ARIC at variant allele fraction (VAF) ³ 2%, which were present in 33,597 (7.7%) UKB participants and 375 (4.9%) ARIC participants. Similar to previous reports, the most commonly mutated genes were *DNMT3A*, *TET2,* and *ASXL1* (Figure 1A, B, Supplementary Figure 1). Mutations in spliceosome genes (*SRSF2*, *SF3B1* and *U2AF1*) were associated with a higher median (25^th^, 75^th^ percentiles) VAF compared to other CHIP mutations in UKB participants (13.0% [7.9%, 24.1%] vs 7.2% [5.0%, 13.7%]; *P* < 0.0001) but not in ARIC (10.1% [7.2%, 20.0%] vs 10.4% [4.9%, 18.2%]; *P* = 0.72) (Figure 1C, D). Most participants with CHIP had only one mutation, with multiple mutations seen in only 8.6% of UKB (n = 2,873) and 12.0% of ARIC participants (n = 45) (Figure 1E, F). As expected, CHIP was associated with older age and smoking status (Table 1).

**Figure 1.**
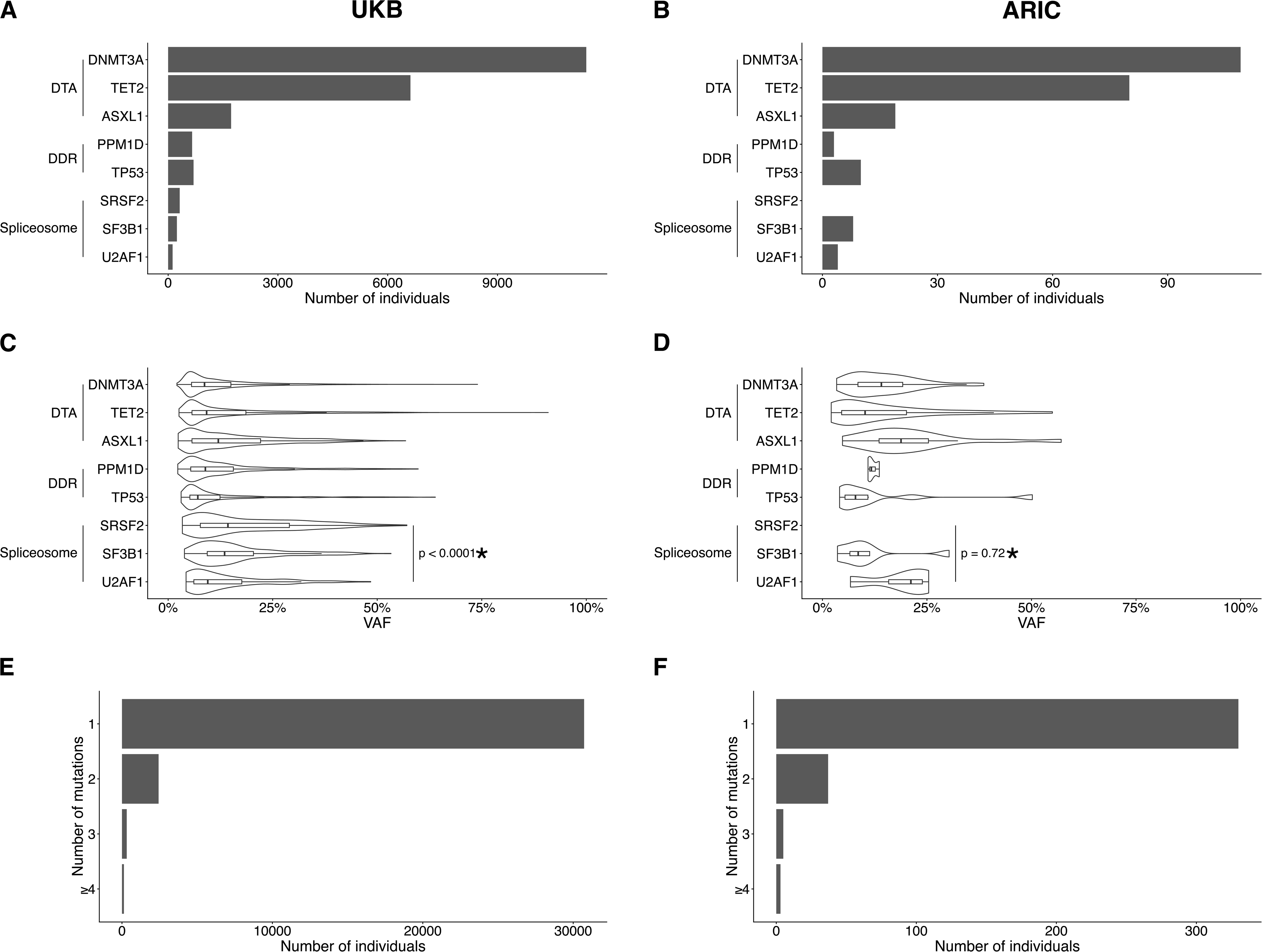
Description of CHIP mutations. Distribution of the number of individuals carrying mutations stratified by genes in **A)** UKB and **B)** ARIC. Distribution of VAF stratified by gene in **C)** UKB and **D)** ARIC. Number of mutations per individual in **E)** UKB and **F)** ARIC. Only the DTA, DDR and classic spliceosome mutations are presented. VAF = variant allele fraction; DTA = *DNMT3A*, *TET2*, *ASXL1*; DDR = DNA damage response. * Denotes the P value for a comparison between spliceosome mutations and other CHIP mutations.

MtDNA heteroplasmy was identified from whole genome sequencing (WGS) data using the MitoHPC pipeline^18^. We used a heteroplasmy VAF threshold of 5% (SNVs between 5% and 95% were called as heteroplasmic) based on extensive simulations and assessment of empirical data to maximize sensitivity to true heteroplasmies while minimizing potential false-positives due to low-level cross-contamination or mis-mapping of nuclear-encoded mitochondrial DNA (NUMTs) ^18^. To identify predicted deleterious mtDNA variants, we used a modified mitochondrial DNA local constraint (mMLC) score, which quantifies the local tolerance to base or amino acid substitution for each base pair in the mtDNA genome ^19^. The mMLC score ranges from 0 to 1, with higher scores indicating more constrained, and therefore, more deleterious SNVs. To capture the overall impact of multiple heteroplasmies in a given individual, we generated the mMLC score sum (mMSS) by summing all mMLC scores for that individual. Prior work has demonstrated that the mMSS is a stronger predictor of overall mortality than heteroplasmy count, suggesting that deleterious mitochondrial heteroplasmies may be causally linked to adverse outcomes ^2^. Heteroplasmy was present in 122,969 (28.3%) and 2,159 (28.3%) participants in the UKB and ARIC, respectively, with heteroplasmies most frequently observed in complex I genes and the D-loop (Figure 2A, B). Heteroplasmic variants in rRNA and tRNA were predicted to be more deleterious than in other mtDNA complex / regions, as reflected by higher mMSS scores (Figure 2C, D). Among those with heteroplasmy, 23,642 (19.2%) participants in UKB and 417 (19.3%) in ARIC had more than 1 heteroplasmy (Figure 2 E, F). Similar to CHIP, heteroplasmy was associated with older age and smoking status (Table 2).

**Figure 2.**
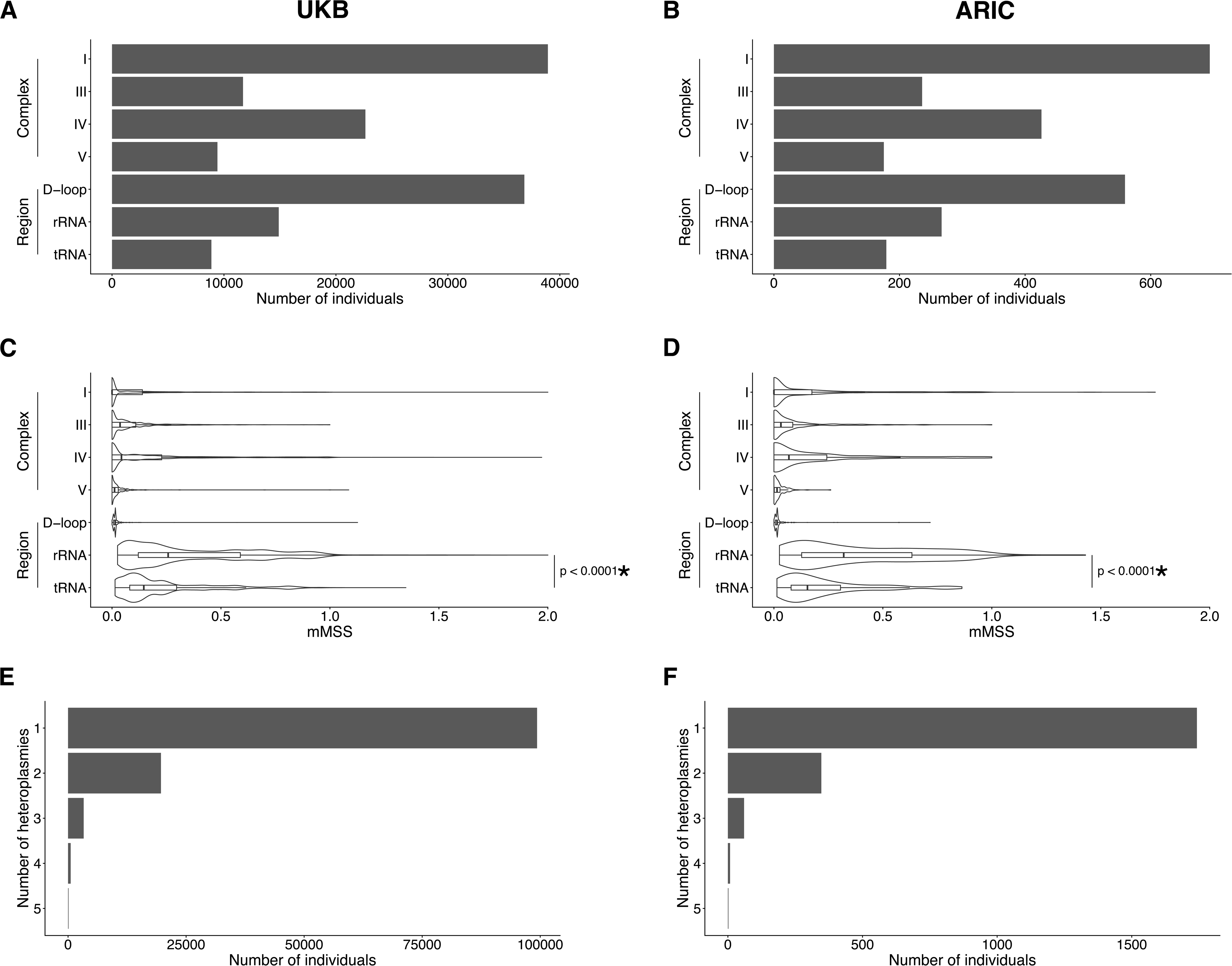
Description of heteroplasmy. Distribution of the number of individuals with heteroplasmy stratified by complex/region in **A)** UKB and **B)** ARIC. Distribution of mMSS stratified by complex/region in **C)** UKB and **D)** ARIC. Number of heteroplasmies per individual in **E)** UKB and **F)** ARIC. * Denotes comparison between rRNA and tRNA mMSS and the mMSS occurring in other complexes/regions.

**Table 2.** Participant characteristics by the presence of heteroplasmy in each cohort.

|  | UK Biobank |  |  | ARIC |  |  |
| --- | --- | --- | --- | --- | --- | --- |
|  | Heteroplasmy<br>(-) | Heteroplasmy | *P* | Heteroplasmy<br>(-) | Heteroplasmy<br>(+) | *P* |
| **Number** | 311,335 | 122,969 |  | 5,473 | 2,159 |  |
| **Age, years** | 56.2 (8.1) | 57.2 (8.0) | <0.0001 | 57.7 (6.0) | 58.3 (6.1) | <0.001 |
| **Men, %** | 45.7 | 46.3 | <0.001 | 44.6 | 46.9 | 0.07 |
| **Race, %** |  |  | <0.0001 |  |  | 0.27 |
| **White** | 94.6 | 95.3 |  | 76.6 | 75.4 |  |
| **Black** | 1.6 | 1.4 |  | 23.4 | 24.6 |  |
| **Asian** | 2.3 | 2.0 |  | 0 | 0 |  |
| **Other** | 1.5 | 1.4 |  | 0 | 0 |  |
| **Ever smoker, %** | 59.3 | 60.8 | <0.0001 | 59.5 | 63.1 | <0.01 |
| **Anemia, %** | 4.2 | 4.3 | 0.43 | 9.5 | 9.3 | 0.89 |
| **Thrombocytopenia, %** | 2.1 | 2.3 | <0.001 | 2.1 | 2.6 | 0.35 |
| **Neutropenia, %** | 1.0 | 1.0 | 0.57 | 2.6 | 4.0 | 0.04 |
| **Cytopenia, %** | 7.0 | 7.2 | 0.07 | 10.6 | 12.8 | 0.08 |
| **MCV, fL** | 91.1 (4.5) | 91.2 (4.6) | <0.0001 | 89.7 (5.1) | 89.9 (4.9) | 0.28 |
| **RDW, %** | 13.5 (1.0) | 13.5 (1.0) | <0.0001 | NA | NA | NA |
| **History of cancer, %** | 6.8 | 7.6 | <0.0001 | 0 | 0 | NA |
| **CHIP (+), %** | 7.1 | 9.3 | <0.0001 | 4.4 | 6.1 | <0.01 |

Continuous variables are displayed as either mean (SD) or median (25th and 75th percentiles). Participant characteristics for both cohorts were based on the time of DNA collection. In ARIC, only participants without a cancer diagnosis at the time of blood collection were included in the analysis. Because of missing blood parameters, in ARIC, we were able to evaluate anemia in 6,473 (84.8%) participants; thrombocytopenia in 5,161 (67.6%) participants; neutropenia in 3,372 (44.2%) participants and cytopenia in 3,344 (43.8%) participants. Abbreviations: ARIC, Atherosclerosis Risk in Communities study; CHIP, clonal hematopoiesis of indeterminant potential; MCV, mean corpuscular volume; mMSS, mitochondrial local constraint (MLC) score sum; NA, not available; and RDW, red cell distribution width.

### Association between CHIP and heteroplasmy

We and others ^2,20–22^ have demonstrated that ∼70% of heteroplasmies identified in adults reflect acquired mutations, and thus, likely mark the clonal expansion of hematopoietic stem cells (HSC). Therefore, we sought to determine whether mtDNA heteroplasmy serves as a more sensitive or alternative marker of CH, or whether it is also involved in biologically relevant processes, contributing to relative expansion of hematopoietic clones either independently or in combination with CHIP mutations. To address this question, we analyzed the association between various metrics of heteroplasmy and CHIP using multivariable logistic regression models adjusted for age, sex, smoking status and a history of cancer. Concurrent CHIP and heteroplasmy (“CHIP and heteroplasmy”) was present in approximately 2-3% of participants (UKB = 2.6%, ARIC = 1.7%) (Figure 3A, B). Heteroplasmy without concurrent CHIP (“Heteroplasmy only”) was seen in approximately 25% of participants (UKB = 25.7%, ARIC = 26.6%), and approximately 5% of individuals had CHIP only without evidence of heteroplasmy (“CHIP only”; UKB = 5.1%, ARIC = 3.2%). Notably, heteroplasmy was more common in CHIP participants compared to those without CHIP in both UKB (34.1% vs 27.8%; *P* < 0.0001) and ARIC (35.2% vs 27.9%; *P* < 0.01) (Figure 3C, D). Among participants with CHIP in UKB, heteroplasmy was more common in individuals with large CHIP clones (VAF ≥ 20%) (44.1% vs 32.2%; *P* < 0.0001) and in those with multiple mutations (39.5% vs 33.6%; *P* < 0.0001). These results were consistent in ARIC, where heteroplasmy was more common in individuals with large clones (42.4% vs 33.1%; *P* = 0.09) and multiple mutations (48.9% vs 33.3%; *P* = 0.04) (Figure 3C, D). Among CHIP genotypes, heteroplasmy was significantly more common in individuals with spliceosome mutations (UKB 55.1% vs 33.6%; *P* < 0.0001 and ARIC 66.7% vs 34.3%; *P* = 0.05) (Figure 3C, D, Supplementary Figure 2).

**Figure 3.**
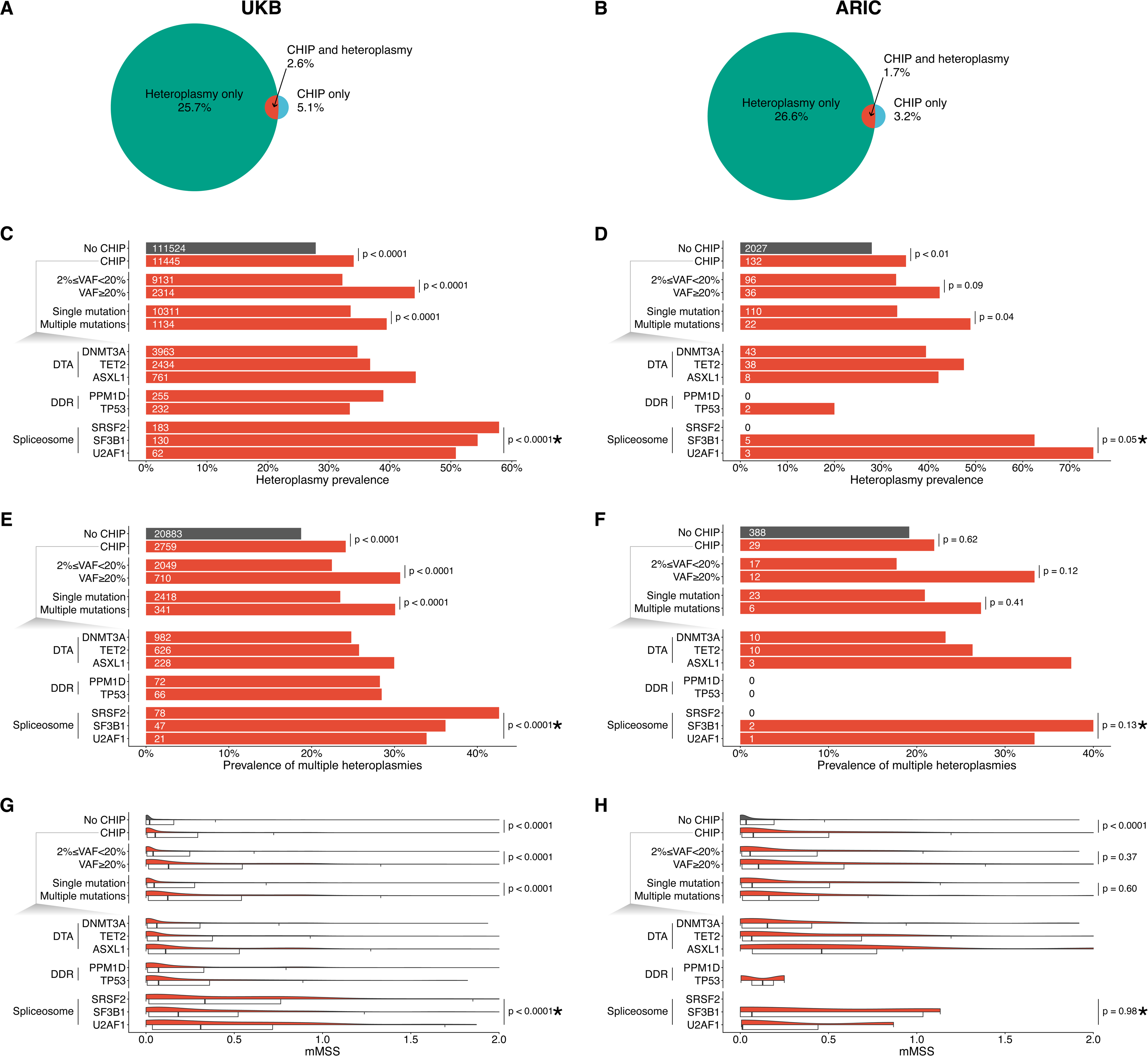
Association between CHIP and heteroplasmy. The percentage of individuals presenting with both CHIP and heteroplasmy, only with CHIP, or only with heteroplasmy in **A)** UKB and **B)** ARIC. The prevalence of heteroplasmy in different CHIP gene subsets is shown in **C)** UKB and **D)** ARIC. The prevalence of multiple heteroplasmies within individuals with heteroplasmy across different CHIP gene subsets is shown in **E)** UKB and **F)** ARIC. The absolute number of individuals with heteroplasmy is indicated by the number on the left side of the bars. mMSS of different CHIP subsets in individuals with heteroplasmy is shown in **G)** UKB and **H)** ARIC. * Indicates the comparison between spliceosome mutations and other CHIP genes. In the UKB, these analyses were adjusted for age modeled as a restricted cubic spline, sex, smoking status and the presence of prevalent cancer. In ARIC, these analyses were adjusted for age modeled as a restricted cubic spline, sex and smoking status. For analyses including mMSS, the models were also adjusted for heteroplasmy count.

Given the association between CHIP and heteroplasmy, we explored whether having multiple heteroplasmies was also associated with features of CHIP in general and, particularly, with high-risk CHIP. Among individuals with heteroplasmy in the UKB, those with CHIP were more likely to have multiple heteroplasmies compared to those without CHIP (24.1% vs 18.7%; *P* < 0.0001) (Figure 3E). CHIP clonal burden (30.7% vs 22.4%; *P* < 0.0001) and multiple CHIP mutations (30.1% vs 23.5%; *P* < 0.0001) were also associated with the presence of multiple heteroplasmies (Figure 3E). Among CHIP genotypes, multiple heteroplasmies were more common in CHIP with spliceosome mutations (39.1% vs 23.6%; *P* < 0.0001) (Figure 3E). Similar results, though not significant, were observed in ARIC, with limited power due to the smaller cohort size (Figure 3F).

To address whether the higher prevalence of heteroplasmy in CHIP individuals was just a marker of CH (passenger mtDNA mutations) rather than likely functionally contributing to clonal expansion, we assessed the association between CHIP and mMSS adjusted for age, sex, smoking status, history of cancer, and heteroplasmy count. Among those with heteroplasmy, the presence of CHIP was associated with a significantly higher mMSS compared to no CHIP (Figure 3G, H, Supplementary Table 2). Moreover, higher mMSS score was observed in individuals with higher clonal burden (VAF ≥ 20%) and with multiple CHIP mutations (Figure 3G, H, Supplementary Table 2). When stratified by specific CHIP genotypes, mutations in spliceosome genes (*SRSF2*, *SF3B1* and *U2AF1*) were associated with a higher mMSS in UKB and had a similar directionality in ARIC when compared to other CHIP mutations (Figure 3G, H, Supplementary Figure 2, Supplementary Table 2). These data suggest that the presence of mtDNA mutations is not merely a marker of clonal expansion but may confer an important cell adaptation.

To determine if the increase in heteroplasmy prevalence in CHIP was due to a specific mtDNA complex / region, we assessed enrichment with measures calculated separately for each complex / region (Supplementary Figure 3). We observed a higher prevalence of rRNA mutations and a lower prevalence of D-loop mutations in participants with CHIP compared to participants without CHIP, and in high-risk subsets of CHIP (VAF>20%, multiple mutations, spliceosome mutations) (Supplementary Figure 3A, B, C, D). Because this enrichment could simply reflect the fact that rRNA mutations are more likely to be deleterious (i.e., heteroplasmies have high average mMLC), we also evaluated the mMSS by complex / region. We observed that, even when restricted to individuals with rRNA mutations, participants with CHIP had higher mMSS values compared to those without CHIP, and, among individuals with CHIP, high-risk subsets of CHIP (VAF>20%, multiple mutations, spliceosome mutations) had higher mMSS values (Supplementary Figure 3E, F, G, H).

### Heteroplasmy and the risk of MN

We have previously reported an association between heteroplasmy and incident and prevalent hematologic malignancies ^2^. Analogous to CHIP and MN, we sought to determine whether the presence of heteroplasmy in the general population was associated with an increased risk of MN. There were 1,191 and 160 cases of incident MN in UKB and ARIC, respectively. Participants with heteroplasmy had a higher risk of developing MN even after adjusting for age, sex, smoking status, and a history of cancer (UKB only) in the UKB (HR = 2.1; 95% CI 1.9–2.3; *P* < 0.0001) and ARIC (HR = 1.7; 95% CI 1.2–2.3; *P* < 0.01) (Figure 4A, B).

**Figure 4.**
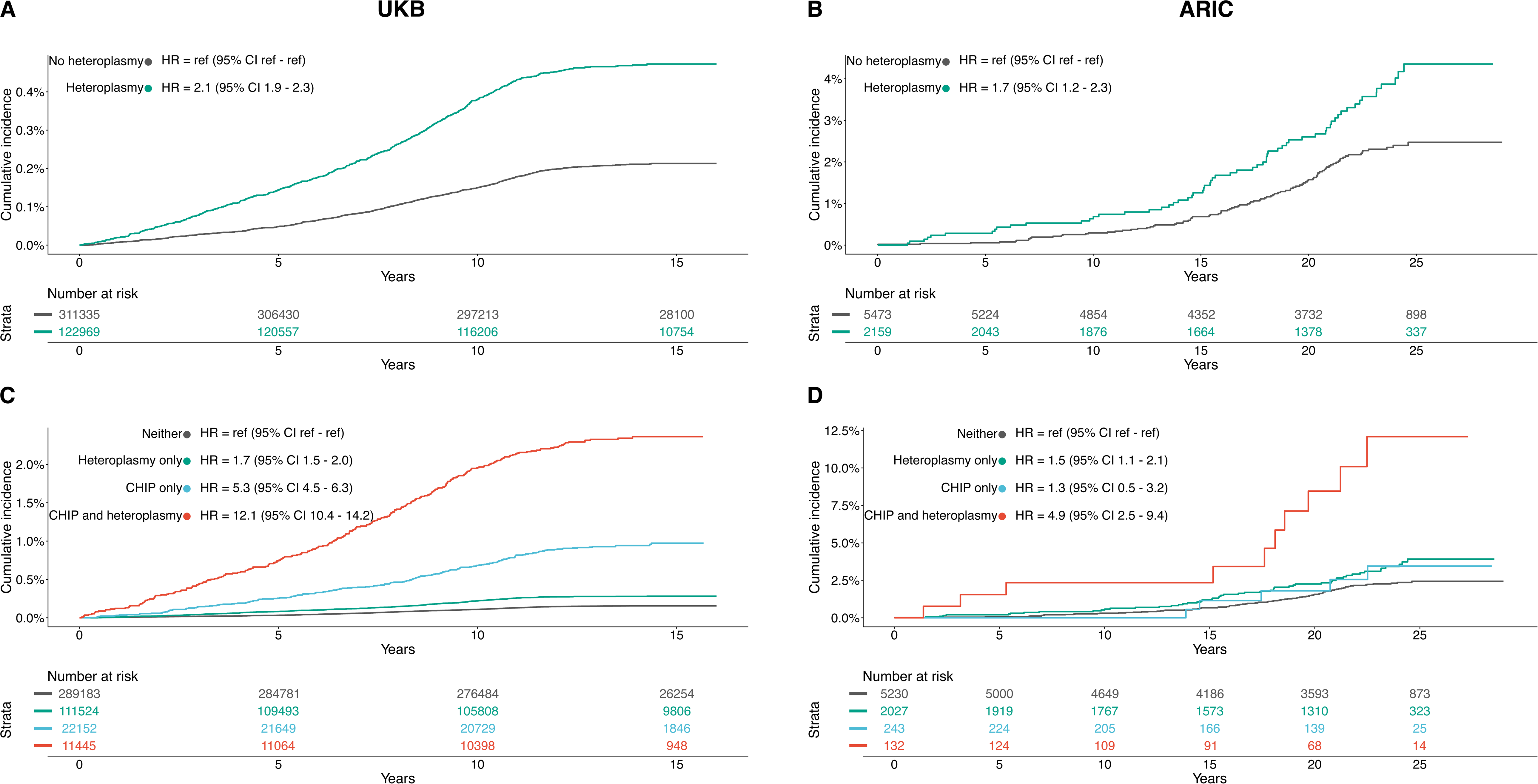
Risk of MN incidence based on CHIP and heteroplasmy status. Kaplan-Meier curves and hazard ratios from the adjusted Cox proportional hazards models comparing the risk of MN development between individuals with heteroplasmy and those without heteroplasmy in **A)** UKB and **B)** ARIC, and those with heteroplasmy only, those with CHIP only and those with both CHIP and heteroplasmy in **C)** UKB and **D)** ARIC. In the UKB, these analyses were adjusted for age modeled as a restricted cubic spline, sex, smoking status and the presence of prevalent cancer. In ARIC, these analyses were adjusted for age modeled as a restricted cubic spline, sex and smoking status.

Among individuals with heteroplasmy, heteroplasmy count was associated with a higher risk of MN in the UKB (HR = 1.7; 95% CI 1.5–1.9; *P* < 0.0001) and ARIC (HR = 1.3; 95% CI 0.9–2.0; *P* = 0.13). Similarly, mMSS was associated with an increased risk of MN in both the UKB (HR = 4.4; 95% CI 3.7–5.2; *P* < 0.0001) (Supplementary Figure 4A, B) and ARIC (HR = 2.2; 95% CI 1.1–4.4; *P* = 0.02). When heteroplasmy count and mMSS were mutually adjusted in the same model, the heteroplasmy count (UKB [HR = 1.2; 95% CI 1.1–1.4; *P* < 0.01] and ARIC [HR =; 95% CI 0.7–1.8; *P* = 0.53]) and mMSS (UKB [HR = 3.7; 95% CI 3.0–4.6; *P* < 0.0001] and ARIC [HR = 2.0; 95% CI 0.9–4.3; *P* = 0.08]) were independently associated with MN, indicating that heteroplasmy is not only a marker for clonal expansion but ostensibly deleterious mtDNA mutations are causally associated with MN. This claim is further supported when analyzing heteroplasmy by complex / region, where D-loop heteroplasmy count, which is largely driven by benign heteroplasmies, showed no association, but D-loop mMSS was significantly associated, with similar effect sizes to the other complexes / regions (Supplementary Figure 5A, B). In the UKB, the associations were independent of race, smoking status and prior cancers. (Supplementary Table 3). Among smokers, the association between heteroplasmy count (HR = 1.2; 95% CI 1.0–1.5; *P* = 0.07), mMSS (HR = 3.2; 95% CI 2.3–4.5; *P* < 0.0001) and MN remained significant after adjusting for the pack-years.

We further explored the potential interaction between CHIP and heteroplasmy in the development of MN by evaluating the associations in individuals with either factor alone and with both CHIP and heteroplasmy. Compared to individuals without either CHIP or heteroplasmy, those with either one alone had an elevated risk of MN (For CHIP, UKB HR = 5.3; 95% CI 4.5–6.3; *P* < 0.0001; ARIC HR = 1.3; 95% CI 0.5–3.2; *P* = 0.55; for heteroplasmy, UKB HR = 1.7; 95% CI 1.5–2.0; *P* < 0.0001; ARIC HR = 1.5; 95% CI 1.1–2.1; *P* = 0.02) (Figure 4C, D). Notably, participants with both CHIP and heteroplasmy (UKB [HR = 12.1; 95% CI 10.4– 14.2; *P* < 0.0001]; ARIC [HR = 4.9; 95% CI 2.5–9.4; *P* < 0.0001]) had a significantly higher risk of MN compared to the expected combined (multiplicative) effect assuming independence of these factors (*P* for interaction = 0.02 in the UKB; *P* for interaction = 0.12 in ARIC). This finding demonstrated a synergistic effect between CHIP and heteroplasmy, potentially amplifying the risk of MN incidence. In the UKB, the association remained significant after adjusting for race, smoking status, and prior cancers (Supplementary Table 3). Finally, we observed that heteroplamsy was an independent predictor of MN risk regardless of the CHIP risk categories when adjusted for the number mutations and clonal burden in UKB (HR = 1.9; 95% CI 1.5–2.3; *P* < 0.0001) and in ARIC (HR = 3.9; 95% CI 1.3–11.8; *P* = 0.02).

### Myeloid malignancy risk score including mitochondrial heteroplasmy in the UK Biobank

A novel clonal hematopoiesis risk score (CHRS) was recently developed in the UKB to better assess the risk of progression to MN among individuals with CHIP^11^. The CHRS is calculated using 8 components, including age ≥65 years, presence of cytopenia, red cell distribution width (RDW) ≥15, mean corpuscular volume (MCV) ≥100, presence of high-risk mutation, a single *DNMT3A* mutation, number of mutations, and VAF ≥20%, and categorized as low-, intermediate-, and high-risk based on 10-year cumulative incidence ^11^. Of the UKB participants with CHIP, 30,542 (90.9%), 2,821 (8.4%), and 234 (0.7%) participants were classified as CHRS low-, intermediate-, and high-risk, respectively, and there were 457 incident cases of MN during a median (IQR) follow-up of 13.7 (12.9, 14.4) years. Compared to those in the low-risk category, intermediate-(HR 10.6; 95% CI 8.6–13.0; *P* < 0.0001) and high-risk (HR = 99.1; 95% CI 76.6–128.3; *P* < 0.0001) categories had a higher risk of MN, after adjusting for sex, smoking status, and a history of cancer (Figure 5). The results were similar when the analysis was restricted to self-reported Whites, unrelated individuals, and never smokers (Supplementary Table 3). In addition, when we performed analysis by subtypes of MN, higher CHRS category was associated with a higher risk of developing all subtypes of MN, including acute myeloid leukemia (AML) and myeloproliferative neoplasms (MPN), and myelodysplastic syndrome (MDS) (Supplementary Figure 6).

**Figure 5.**
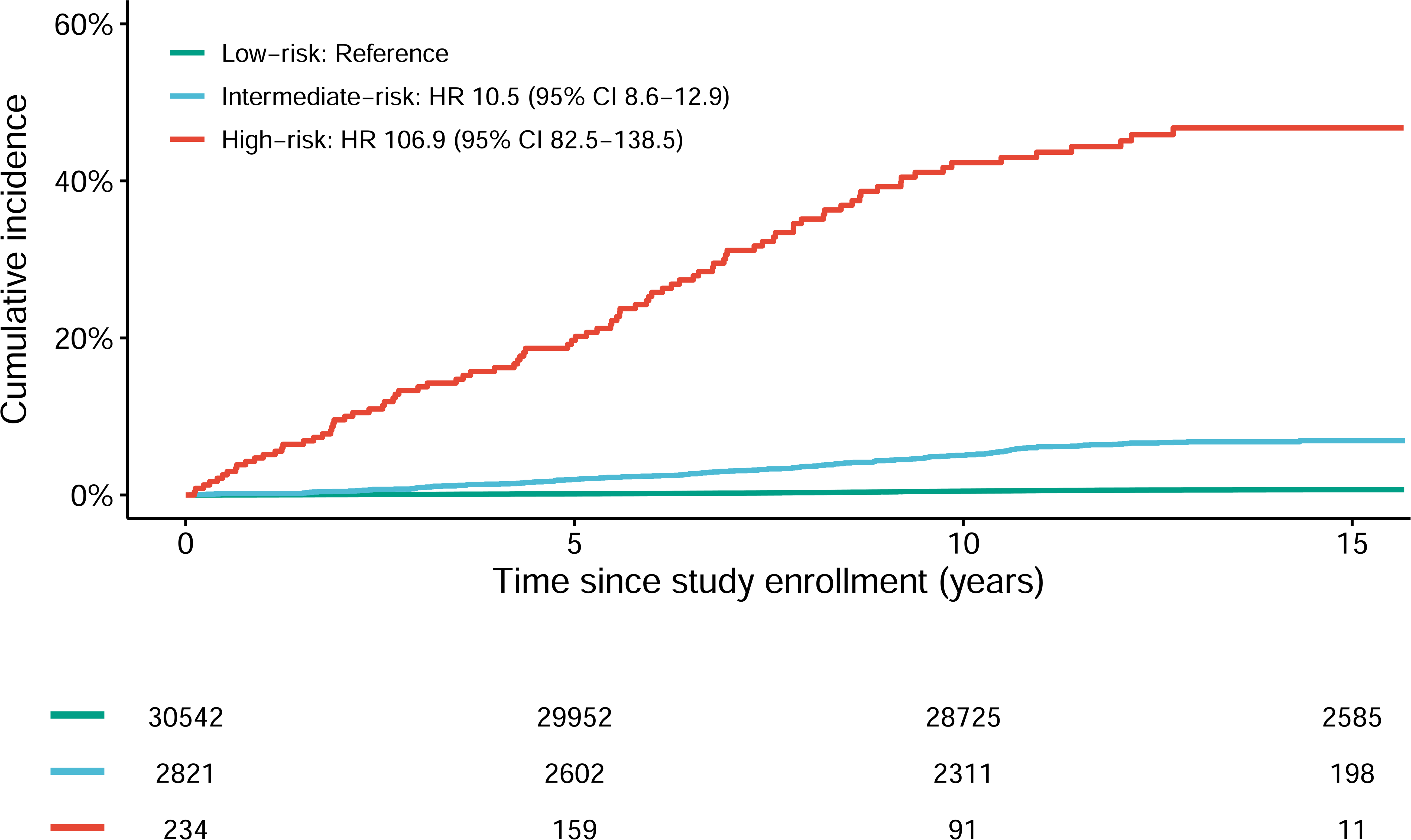
Cumulative incidence and hazard ratios (95% confidence intervals) of MN by CHRS category. Kaplan-Meier curves and hazard ratios from the adjusted Cox proportional hazards models comparing the risk of MN development by CHRS category. The analysis was adjusted for age modeled as a restricted cubic spline, sex, smoking status and the presence of prevalent cancer. Abbreviations: CI, confidence interval; and HR, hazard ratio. Green line indicates low-risk, blue line indicates intermediate-risk, and red line indicates high-risk groups.

Compared to those in the low-risk CHRS category, participants in the intermediate– and high-risk categories were, on average, more likely to have a higher heteroplasmy count and mMSS (Supplementary Table 4, Supplementary Figure 7). We further evaluated whether heteroplasmy was associated with MN independent of CHRS. Both heteroplasmy count (HR 1.4; 95% CI 1.3–1.6; *P* < 0.0001) and mMSS (HR 2.1; 95% CI 1.7–2.5; *P* < 0.0001) were associated with incident MN, after adjusting for CHRS category, sex, smoking, and a history of cancer. When heteroplasmy count and mMSS were mutually adjusted for in the same regression model, the associations were attenuated (HR 1.3; 95% CI 1.1–1.4; *P* = 0.001 for heteroplasmy count and HR 1.5; 95% CI 1.1– 2.0; *P* = 0.007 for mMSS) but retained their significance, suggesting that heteroplasmy is not only a biomarker, but deleterious variants are also causally associated with MN.

Having established that heteroplasmy is a predictor of MN independent of CHRS, we updated the CHRS model (CHRS-M) to incorporate the presence of heteroplasmy and mMSS, using the algorithm as outlined in the original CHRS manuscript (round the effect estimate to the nearest 0.5 and add 1) ^11^ (Table 3, Supplementary Figure 8). To account for the two additional heteroplasmy parameters in the score (presence of heteroplasmy and mMSS), we added 2 to the score cutoffs used by the CHRS to classify risk groups; low (9.5–12.5; n = 29,147), intermediate (13–15; n = 4,089), and high (15.5–18.5; n = 361) (Figure 6A). The cumulative incidence of MN at 10 years was 0.4%, 3.1%, and 30.7% in each risk group, respectively (Figure 6B, C). Compared to the low-risk group, the intermediate– and high-risk groups had an increased risk of incident MN (HR 7.2; 95% CI 5.8–9.0; *P* < 0.0001; and HR 87.1; 95% CI 68.8– 110.6; *P* < 0.0001, respectively) after adjusting for sex, smoking status, and a history of cancer. Compared to the CHRS, CHRS-M resulted in reclassification of 1,395 individuals (4.6%, 22 cases) from low to intermediate risk and 127 (4.5%, 31 cases) from intermediate to high risk, resulting in a 34.4% increase of incident cases identified as high-risk individuals (n = 90 using CHRS vs. n = 121 using CHRS-M). Including parameters for heteroplasmy in the prediction model significantly improved the discrimination assessed by net reclassification index (NRI 8.7; 95% CI 5.4–11.7; NRI in cases 13.1; 95% CI 9.9–16.2; NRI in controls –4.4; 95% CI –4.6–-4.2), indicating that adding heteroplasmy information to the existing CHRS score improves the sensitivity of identifying those who develop MN, and particularly those are at a high risk, with a small decrease in specificity.

**Figure 6.**
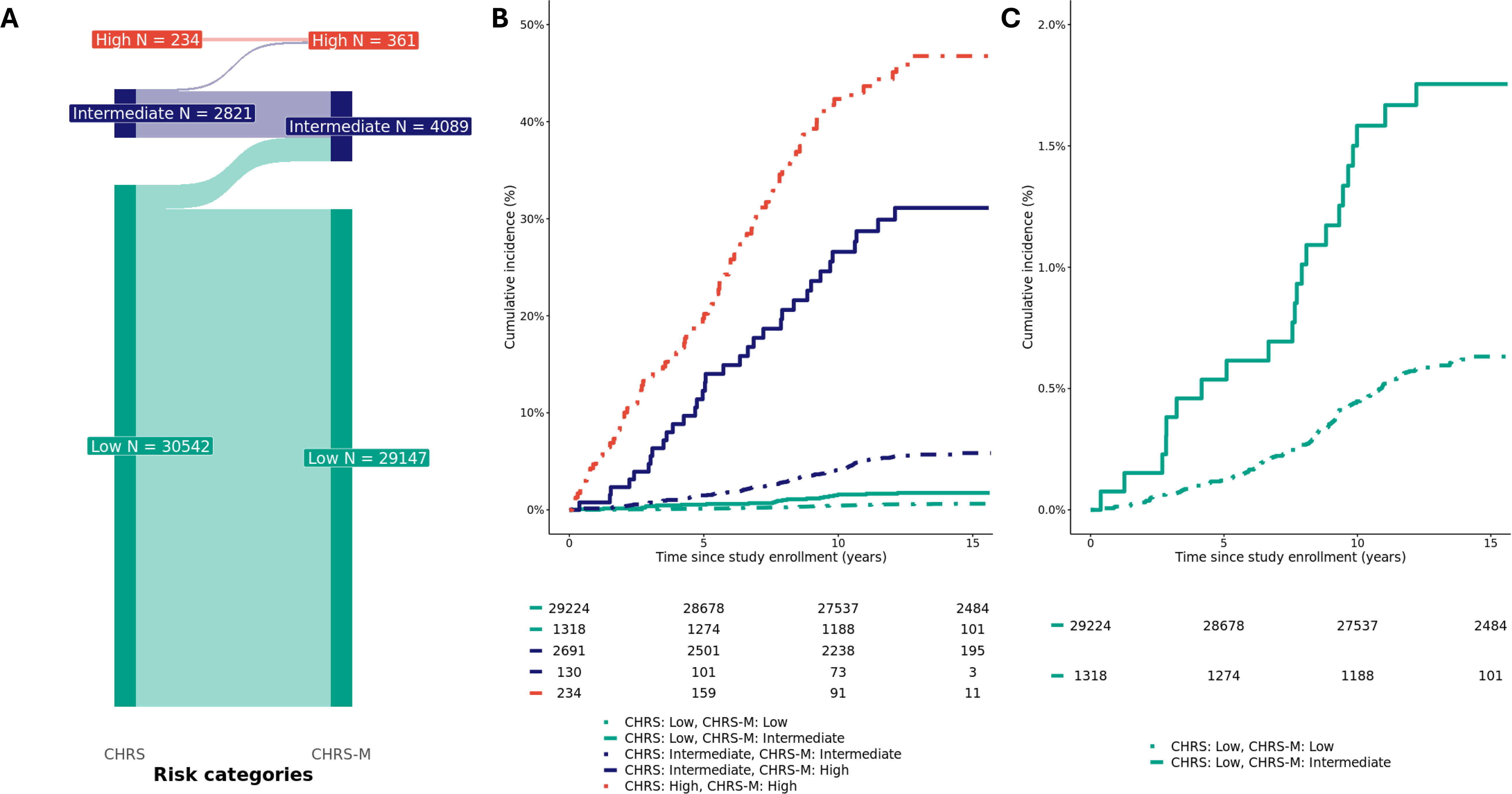
Risk of MN development by CHRS-M categories. (**A**) A Sankey diagram of the reclassification of CHRS to CHRS-M. (**B**) Kaplan-Meier curves for the risk of MN development by CHRS-M category. The dotted lines indicate the cumulative incidence of individuals who remain in the same risk category. The solid lines indicate the cumulative individuals who are recategorized from low to intermediate (green) risk and from intermediate to high (dark blue) risk categories using the CHRS-M. (**C**) Kaplan-Meier curves for the risk of MN development in the CHRS low-risk group with the same data as (**B**) on an enlarged y axis. Abbreviations: CHRS, clonal hematopoiesis risk score; and CHRS-M, clonal hematopoiesis risk score with mitochondrial heteroplasmy.

**Table 3.** Values assigned to component variables of the CHRS modified for heteroplasmy ^11^.

| **Variables** | **0.5** | **1** | **1.5** | **2** | **2.5** |
| --- | --- | --- | --- | --- | --- |
| **Age (years)** |  | <65 | ≥65 |  |  |
| **Cytopenia** |  | CHIP | CCUS |  |  |
| **RDW** |  | <15 |  |  | ≥15 |
| **MCV** |  | <100 |  |  | ≥100 |
| **High-risk mutation** |  | Absent |  |  | Present |
| **Single *DNMT3A*** | Present | Absent |  |  |  |
| **Number of mutations** |  | 1 |  | ≥2 |  |
| **VAF** |  | <20% |  | ≥20% |  |
| **Heteroplasmy** |  | Absent | Present |  |  |
| **mMSS** |  | < 0.59 | 0.59 – 1.78 | ≥1.78 |  |

Abbreviations: CCUS, clonal cytopenia of undetermined significance; CHIP, clonal hematopoiesis of indeterminant potential; MCV, mean corpuscular volume; mMSS, mitochondrial local constraint (MLC) score sum; RDW, red cell distribution width; and VAF, variant allele fraction.

### The role of heteroplasmy on myeloid malignancy in individuals without CHIP

We further evaluated the association between heteroplasmy and MN in individuals without CHIP in the UKB (n = 400,707). Among individuals without CHIP, there were 734 incident cases of MN during a median (IQR) follow-up of 13.8 (13.1, 14.5) years. Both the number of heteroplasmies and mMSS were associated with a higher risk of MN (HR = 1.5; 95% CI 1.4–1.6; *P* < 0.0001 for a 1-unit increase in heteroplasmy count and HR = 3.1; 95% CI 2.5–3.8; *P* < 0.0001 for a 1-unit increase in mMSS) when adjusted for age, sex, smoking status, and a history of cancer. The associations were attenuated when we assessed the independent associations of heteroplasmy count (HR = 1.2; 95% CI 1.1–1.4; *P* < 0.01) and mMSS (HR = 2.3; 95% CI 1.7–3.1; *P* < 0.0001) with MN by mutually adjusting for each other. Heteroplasmy count (HR = 1.2; 95% CI 1.1–1.4; *P* < 0.01) and mMSS (HR = 2.2; 95% CI 1.6–3.0; *P* < 0.0001) were independent risk factors of MN even after further adjusting for blood profiles that are potential biomarkers of MN, including RDW, MCV, and presence of cytopenia, suggesting that mtDNA heteroplasmy is a novel predictor of MN risk also in people without CHIP.

## Discussion

The role of somatic nuclear DNA mutation, and CHIP in particular, has been increasingly recognized as a key risk factor for developing MN. Much less is known about the role of somatic mutations in the mitochondrial genome, with recent work identifying a strong association between mitochondrial heteroplamsy and hematological cancers ^2^, and the interaction between nuclear and mitochondrial mutations ^23^. In this study, we found a significant enrichment of participants harboring both CHIP variants and mitochondrial heteroplasmy, with compelling evidence that this enrichment differs based on the specific natures of the nuclear and mitochondrial mutations. We further found that mitochondrial heteroplasmy can significantly improve a CHIP-based risk score for the development of MN and may help identify those at risk in non-CHIP individuals, in whom no current risk scores are currently available.

The high prevalence of heteroplasmy (approximately 30% in both UKB and ARIC), and the observation that the majority of heteroplasmies (70%) measured in peripheral blood are somatic, suggest that they can serve as markers of clonal expansion ^2,20–22^. This observation is corroborated by the higher prevalence of heteroplasmy in individuals with CHIP, particularly with VAF ≥20%, compared to those without CHIP. Somatic mitochondrial mutations have also been demonstrated to play an important role in tumorigenesis by increasing the level of reactive oxygen species (ROS) and reducing apoptosis ^24–26^. Moreover, certain tumors have been found to significantly favor the selection of deleterious mitochondrial mutations ^24,27^. Consistent with these results, we found that heteroplasmic variants predicted to be more deleterious were enriched in individuals with high-risk CHIP, particularly in individuals with significantly expanded clones (VAF ≥20%), with more than one mutation, and with mutations in spliceosome machinery ^7,11,28–30^. These associations remained significant even after adjusting for the number of heteroplasmies, supporting the claim that the functional nature of the mitochondrial mutations is important, and does not simply represent passenger mutations due to CHIP.

An important aim of our study was to determine whether the presence of heteroplasmy was merely a surrogate marker for CH or whether it played a causal role in the development of MN. By incorporating both heteroplasmy presence (count) and a predicted burden metric (mMSS) in the same regression model, and demonstrating that both remain significant predictors, we provide strong statistical evidence for a causal role of deleterious mtDNA variation in MN pathogenesis. This causal role is further corroborated by our finding that there is a statistically significant synergistic (non-multiplicative) effect of heteroplasmy and CHIP on the risk of MN. Thus, beyond simply serving as a marker of clonal hematopoiesis, the specific functional consequence of the heteroplasmy itself likely plays a role in tumorgenesis, potentially including oxidative damage to oncogenes or tumor suppressor genes and leading to abnormal hematopoiesis ^31^. Indeed, heteroplasmy has been found to be an adverse prognostic factor in patients diagnosed with MN ^32–34^.

From a clinical perspective, the ability to identify high-risk MN individuals early is key to improving outcomes. Incorporating heteroplasmy information into a state-of-the-art CHIP-based prediction model significantly improved the sensitivity of identifying individuals at risk of developing MN, with little loss of specificity. Specifically, integrating the presence (heteroplasmy yes/no) and predicted deleterious effect of heteroplasmic variants, the updated model was able to detect 34.4% more incident cases among individuals classified as high-risk (10-year risk ≥ 10%) of developing MN. This is a remarkable improvement to the current score system, without the need for recalibrating or retraining the algorithm. Clinically, this finding suggests that heteroplasmy information can refine the existing risk stratification and identify high-risk individuals (10-year risk ≥10%) who have been inappropriately classified as intermediate risk (CHRS 10-year risk of 2-10%), and who may benefit from close surveillance. Moreover, mitochondrial DNA sequencing and detection of heteroplasmy can be feasibly implemented in currently used molecular tests to facilitate the identification of this high-risk population.

While CHIP has great clinical utility, particularly when combined with heteroplasmy, the risk of MN is particularly not well understood among individuals without evidence of CHIP. Our study demonstrates that having any mitochondrial heteroplasmy was associated with a higher risk of MN, even after accounting for known risk factors in individuals without CHIP. Notably, higher mMSS was a more robust predictor for MN than just the number of heteroplasmies, further suggesting that mutation in mtDNA may be an independent causal mechanism for MN. However, MN is a rare disease, with a crude incidence rate of 14 cases per 100,000 person-years in the general population ^35^, making it particularly difficult to precisely estimate its risk and study novel biomarkers. Consequently, a risk stratification model applicable to the general population is currently unavailable. Further studies are needed to better understand the underlying mechanisms linking mtDNA variation to MN incidence and to develop a universal risk model for MN to identify high risk population regardless of CHIP status.

The current manuscript has several limitations. First, as there are differences between UKB and ARIC and between each cohort and the population from which they were sampled. Nonetheless, the similarity of the results between the two cohorts only strengthens the validity of our observations. Secondly, the use of WES to detect CHIP has inherent limitations related to the relatively shallow sequencing depth, which can lead to an increase in false negatives. In addition, the UKB is mostly of self-reported White individuals and the subgroup analysis of non-White individuals was based on a small number of events (34 events out of 23,991 individuals). Thus, the findings of the interaction between heteroplasmy and CHIP, as well as the CHRS-M need to be validated in more diverse populations. Despite these limitations, our findings suggest that heteroplasmy, in addition to being a marker of clonal expansion, may be a causal biomarker of MN development, with clinical utility in the general population.

## Methods

### Study population

The UK Biobank (UKB) is a large population-based prospective study of ∼500,000 participants between the ages of 40 to 69 years across the United Kingdom recruited from 2006 to 2010 ^36^. The UKB has extensive information on participant’s genetic and phenotypic data, including demographics and lifestyle factors. The data is linked to the death registry, cancer registry, hospital admissions, and primary care visit data.

For the current study, 434,404 participants who had both WGS, which was used for calling mitochondrial DNA heteroplasmy, and WES, which was used for CHIP variant calling, and passed the variant-level and sample-level QC for both and met inclusion / exclusion criteria were analyzed (details below; Supplementary Figure 9). More specifically, there were 490,355 participants with WGS, where mtDNA heteroplasmy was evaluated using MitoHPC ^18^ (https://github.com/dpuiu/MitoHPC). We excluded samples (n = 3,776) if they met one of the following criteria: suspicious of having 1) potential mitochondrial contamination (a contamination level ≥3% from Haplocheck ^37^), 2) 2 or more variants belonging to a different mitochondrial haplogroup, 3) 2 or more variants matching to the same NUMT, or 4) low minimum base coverage (<100) or low mean base coverage (<500). In addition, because the presence of NUMTs can influence the false positive mtDNA heteroplasmy calls at low mtDNA-CN ^38^, we removed 12,001 participants with mtDNA-CN ≤40. We additionally removed 704 participants with a heteroplasmy count above 5 because we found that samples with a heteroplasmic count above 5 are not distinguishable from contaminated samples ^2^. Some samples met multiple exclusion criteria, which resulted in 13,921 participants being excluded. After exclusion, there were 476,434 participants with heteroplasmy information. For WES, 466,042 participants had both CHIP calls from GATK Mutect2 ^39^ and *U2AF1* calls. No additional participants were excluded for having a high number of INDELs or CHIP variants. Of the 450,916 participants who passed QC for heteroplsmy and CHIP, we excluded 463 participants with a history of MN (n = 310) or a potential MPN (n = 182). We further excluded 16,149 participants who did not have information on tobacco smoking status (n = 2,211) or have missing values for any one of the following measurements that are used for calculating the clonal hematopoiesis risk score (CHRS; n = 14,077): hemoglobin, platelet count, neutrophil count, red blood cell distribution width (RDW), or mean corpuscular volume (MCV). A detailed description of QC steps for heteroplasmy and CHIP, and the exclusion for heteroplasmy count and potential MPN are provided below. The final sample for analysis included 434,404 individuals (199,046 men and 235,258 women). The current study was approved by the Johns Hopkins Medicine Institutional Review Boards.

The Atherosclerosis Risk in Communities (ARIC) study is a community-based, prospective cohort study focusing on the risk factors for cardiovascular disease, that recruited 15,792 individuals between the ages of 45 and 64 from 4 communities in the US (Forsyth County, NC; Jackson, MS; Minneapolis suburbs, MN; and Washington County, MD) from 1987 to 1989 ^40^. Of those, 12,776 had WGS available, on which MitoHPC was run (Supplementary Figure 10). After excluding participants that failed heteroplasmy QC and those that did not have visit information (n = 69), we retained 12,707 individuals with heteroplasmy information. We further excluded individuals with those that did not have WES data from the same visit (n = 4,340), that did not have information on incident myeloid neoplasms (n = 709), that were likely prevalent MPNs (n = 0), that had an excessive number (≥4) of CHIP indels (n = 3) and that had 10 or more CHIP mutations (n = 1) and did not have information on smoking status (n = 22). The final sample for analysis included 7,632 individuals (3,453 men and 4,179 women). All participants included in this study provided appropriate informed consent. The ARIC study protocol was approved by the Johns Hopkins Medicine Institutional Review Board (IRB), which serves as the single site IRB.

### Heteroplasmy and mitochondrial DNA copy number analysis in the UK Biobank

Samples for DNA were collected on the baseline visit and DNA was extracted from buffy coat using the Maxwell® 16 Instrument (Promega) and the Maxwell® 16 Blood DNA Purification Kit (Promega – AS1010X). WGS CRAM files from the UKB were processed on the DNA Nexus server. For heteroplasmy identification, we used the MitoHPC pipeline (version 20230418; all default settings with default random down-sampling to use at most 222K reads) ^18^, implementing GATK Mutect2 for variant identification ^39,41^. We defined a heteroplasmic SNVs at a variant allele frequency of 5%, meaning that variant alleles at a frequency of 5-95% within an individual are defined as heteroplasmic. Alleles less than 5% or greater than 95% are counted as homoplasmic. To test the robustness of this cutoff, we repeated the main analyses with VAF cutoffs at 3% and 10%. The results were consistent across different cutoffs (Supplementary Figure 11). MitoHPC pipeline incorporates haplogrep (https://github.com/seppinho/haplogrep-cmd/v2.4.0) ^42^ to identify haplogroups and Haplocheck (https://github.com/genepi/haplocheck) ^37^ to detect in-sample contamination by detecting two different mitochondrial haplotypes in each sample. For mitochondrial DNA copy number (mtDNA-CN) calculation, we used SAMtools ^43^ embedded in MitoHPC to generate read count and coverage information, using the command ‘samtools idxstats’. A detailed documentation on how to run MitoHPC on DNA Nexus server is available: https://github.com/ArkingLab/MitoHPC/blob/main/docs/DNAnexus_CLOUD.md.

All 490,355 WGS samples processed using MitoHPC variant calling were successfully completed ^18^. The mean nuclear genomic coverage for WGS samples in the UKB was 98x and the mean mtDNA coverage was 965x.

### Heteroplasmy and mitochondrial DNA copy number analysis in ARIC

We analyzed WGS data from the Atherosclerosis Risk in Communities (ARIC) study (n = 12,776). DNA samples were collected for each participant across multiple clinic visits (V1, V2, V3, V4, MRI and V5) and DNA for WGS were isolated from buffy coat using the Gentra Puregene Blood Kit (Qiagen). WGS calls were from the Trans-Omics for Precision Medicine (TOPMed; https://topmed.nhlbi.nih.gov/methods) program are from freeze 8 (30.6%; n = 3,915) and from the Centers for Common Disease Genomics (CCDG; https://www.genome.gov/27563570) initiative (69.4%; n = 8,861). There were 11 individuals who were sequenced in both TOPMed and CCDG, and we randomly selected one of the two. TOPMed studies provide WGS data at ∼30x genomic coverage using Illumina next-generation sequencing technology, which must pass specific quality control metrics before being released for use by the scientific community. The median mtDNA coverage was 1898x.

### Heteroplasmy filtering for variants

MitoHPC provides various variant annotations to evaluate variant quality, allowing us to remove low quality variants prior to analysis. We excluded variants with read depth <300 and those flagged as base quality, mapping quality, strand bias, slippage, weak evidence, position, clustered, fragment length, and haplotype flags in the FILTER column of the VCF. We further excluded heteroplasmic variants at poly-C homopolymer regions on the mitochondrial chromosome (the list is provided as HP.bed.gz in https://github.com/dpuiu/MitoHPC). INDELs are also excluded because they are often found in homopolymer regions, where it is challenging to accurately call heteroplasmies ^18^.

### Mitochondrial local constraint (MLC) score

The mitochondrial local constraint (MLC) score is a metric reflecting local tolerance to substitutions of base or amino acid. It is calculated for every possible mtDNA single nucleotide variant (SNV) by applying a sliding window method ^19^. In brief, starting from position m.1, within a window of 30 bases, the observed:expected (oe) ratio of substitutions and its 90% confidence interval (CI) was calculated in gnomAD ^38^. The window start position is then moved by 1 bp, and this process is iterated until the full length of mtDNA is covered (i.e., a start position of m.16569). For positions in protein genes, calculations are restricted to missense variants, or substitutions in amino acids, while for all other positions, all base substitutions are used. The mean of oe ratio 90% CI upper bound fraction (OEUF) is calculated for each position using all overlapping windows, and then percentile ranked to achieve a positional score between 0 and 1, where 1 is most constrained and 0 is least constrained. An MLC score is obtained for every mtDNA SNV as follows: non-coding, RNA, and missense variants are assigned their positional score; and non-missense variants in protein genes are assigned scores based on the OEUF value of the variant class, with synonymous, stop gain, and start/stop lost being assigned scores of 0.0, 1.0, and 0.70, respectively. Variants with higher scores are predicted to be more deleterious. To account for heteroplasmic variants that also present as homoplasmic in the population, we refined the MLC score to account for the frequency of homoplasy in the UKB population by calculating modified MLC (mMLC) as MLC/(1 + log_10_(number of individuals with homoplasmy in the UKB + 1)). mMLC for both UKB and ARIC was calculated using the individuals with a given homoplasmic variant in the UKB. To capture the functional impact of multiple heteroplasmies, we calculated an mMLC score sum (mMSS) by summing all mMLC scores within a given individual.

### CHIP variant calling

WES CRAM files were aligned to hg38 and hg19 in the UKB and ARIC, respectively. Variant calling was performed using Genome Analysis Toolkit (GATK) v.4.2.2. Mutect2 ^39,44^. Mutect2 was run in ‘tumor-only’ mode using non-default parameters: gatk Mutect2 … –-panel-of-normals ${ref_pon} –-germline-resource ${ref_germ}. Raw variants called by Mutect2 were filtered out with FilterMutectCalls using the estimated prior probability of a reading orientation artifact generated by LearnReadOrientationModel.

### U2AF1 calling in UKB (hg38)

Considering the known duplication of *U2AF1* in hg38, we called these variants using a previously described pileup approach (https://github.com/weinstockj/pileup_region) ^45^. Briefly, reads containing *U2AF1* pathogenic variants and the total number of reads at the given locus were counted at both the *U2AF1* and *U2AF1L5* loci. The alternative depth and total depth were considered as the average between the two loci. VAF was calculated as the alternative depth divided by total depth and then multiplied by 100.

### CHIP variant filtering

Variants were annotated using ANNOVAR ^46^ and filtered based on: 1) the presence in a gene part of a custom CHIP panel (Supplementary Table 1); 2) a previously published whitelist ^45^; 3) Mutect2 FILTER of PASS, weak_evidence, or germline; 4) depth ≥20; 5) alternate allele count ≥3 in the case of SNVs; 6) alternate allele count ≥5 in the case of MNVs; 7) ALT F1R2 ≥1; 8) ALT F2R1 ≥1; 9) VAF ≥2% in the case of SNVs; 10) VAF ≥10% in the case of indels; 11) exclusion of synonymous SNVs; 12) the maximum allele frequency across non-cancer populations (non_cancer_AF_popmax) under 0.001 in the gnomAD exome collection v2.1.1 ^47^; or 13) recurrent germline variants considered as such based on their presence in 3 or more individuals and having a binomial test p-value ≥0.01 in ≥80% of those individuals. Variants occurring on genes located on the X chromosome in males had their VAF divided by 2. For ARIC WES, given the higher noise, an alternate allele count ≥5 was used for SNVs. The higher noise seen in ARIC was reflected through the higher number of SNVs seen in the VCF per individual compared to UKB.

On a sample level, we excluded individuals with more than 3 mutations, all of which are represented by MNVs, individuals with more than 9 CHIP variants, or individuals with suspected of having potential MPNs, as described in Supplementary Figures 9 and 10.

Large CHIP clones were defined as those having a VAF ≥20%. Small CHIP clones were defined as having a VAF ≥2% and <20%. Potential MPNs were defined as individuals with known pathogenic variants in *JAK2*, *CALR,* or *MPL,* and had a hematocrit over 48% or a platelet count over 450×10^3^/µL. If either of these laboratory parameters were missing, the individual was excluded. Of note, although *CALR* was not included in the CHIP panel, it was assessed because *CALR* is a recurrently mutated gene in MPN. The same filtering steps were applied for identifying *CALR* mutation. However, the presence of *CALR* mutation was not used to define CHIP in the downstream analysis.

### Definition of MN

In the UK Biobank, myeloid neoplasm (MN) and its subtypes were defined using ICD-9 and ICD-10 codes from the cancer registry linked to the UK Biobank (Supplementary Table 5)^11^. More specifically, MN was defined as the presence of acute myeloid leukemia (AML), chronic myeloproliferative disease / myeloproliferative neoplasms (MPN), myelodysplastic syndrome (MDS), or chronic and other myelogenous leukemia.

In ARIC, hematological malignancy (HM) cases were ascertained through 12/31/2015 via linkage with cancer registries in the four states where the ARIC participants were recruited, and supplemented with medical records, routinely collected hospital discharge summaries, and death certificates ^48^. MDS before 2001 and MPN before 2011, both characterized by clonal expansion, were considered to be pre-leukemia; today, these are considered to be leukemia. These pre-leukemias were not captured by cancer registries before those dates. Thus, we used 2 strategies to identify MDS and MPN: 1) identified cases using ICD codes from routinely collected hospital discharge summaries, 2) identified cases using ICD codes from CMS claims data (Supplementary Table 6). Participants with an ICD code consistent with MDS, MPN, or other such state prior to or concurrent with the blood sample used to call CHIP from WES were excluded. Complete blood cell counts are measured as part of the ARIC protocol but were not available for all participants at their WES visit. For the participants with an eligible MDS or MPN ICD code, we reviewed their complete blood cell counts (if available) that were measured in the same blood sample used to call CHIP. We excluded as cases any participant with blood count anomalies suggestive of an undiagnosed HM. Using the histology codes from the cancer registries and ICD codes from the hospital discharge summaries and claims data, we classified the cases as myeloid-or lymphoid-derived cases. Three experts in hematologic malignancies and epidemiology (L.P.G, M.R., S.P.) adjudicated all of these cases.

### Statistical analysis

Categorical data was represented as absolute count (percent). Frequencies of two categorical variables were analyzed using Fisher’s exact test. Continuous variables are presented as mean (SD) for variables with Gaussian distribution or median (25^th^, 75^th^ percentiles) for variables with non-Gaussian distribution. Normality of the distribution was assessed using skewness, kurtosis and histogram visualization. Differences between two groups with non-Gaussian distribution were assessed using the Mann-Whitney-Wilcoxon rank sum test. Differences between two groups for variables with Gaussian distribution were assessed using Student’s t-test. We used multivariable logistic regression models to evaluate the associations between CHIP and a binary variable for heteroplasmy. For the association between CHIP and mMSS, we used multivariable linear regression models. If the dependent variable was right skewed, we added 1 and log-transformed it. For UKB, these analyses were adjusted for age modeled as a restricted cubic spline (with 4 degrees of freedom), sex, tobacco smoking status (never or ever), and a history of cancer. For ARIC, the analyses were adjusted for age modeled as a restricted cubic spline (with 4 degrees of freedom), sex, and cigarette smoking status (never or ever). History of cancer was not included as a covariate in ARIC as the follow-up duration since the WGS / WES visit was only available for those who did not have cancer prior to or at the WGS / WES visit.

For survival analysis, we used time from DNA collection to the development of MN, death, or end of follow-up (administrative censoring on December 22, 2022 in the UKB, and December 31, 2015 in ARIC), whichever occurred first. Cumulative incidence of MN was presented as Kaplan-Meier curves. Hazard ratios and corresponding 95% confidence intervals were estimated using a multivariable Cox proportional hazards model, adjusting for age (restricted cubic splines with 4 degrees of freedom), sex, tobacco smoking (never or ever), and a history of cancer. In ARIC, as the analysis was restricted to individuals without a history of cancer, adjustment was made for age at the time of DNA collection (restricted cubic splines with 4 degrees of freedom), sex, and cigarette smoking status (never or ever) at the time of DNA collection.

We performed several sensitivity analyses to demonstrate the robustness of findings. Due to the limited number of events in ARIC, sensitivity analyses were only performed in UKB. To address the potential differences in the association between mMSS and health outcomes by race/ethnicity ^2^, we performed an analysis restricted to self-reported White (N = 410,313) and non-White (N = 23,991) individuals, separately. As the UKB includes related individuals, we repeated the analysis restricted to genetically unrelated individuals (N = 362,132; defined as variable “used.in.pca.calculation” as provided by the UKB). In addition, because adjusting for smoking status simply as a binary variable may be subject to residual confounding ^49^, we further adjusted for pack-years of smoking in an analysis using former or current smokers (N = 131,562). We additionally performed an analysis restricted to never smokers (N = 238,086). Furthermore, we evaluated the association of heteroplasmy, CHIP, and subtypes of MN (AML, MDS, and MPN), separately. Finally, we repeated the analysis after excluding participants with a history of cancer (N = 403,732). For this analysis, adjustment was made for age (restricted cubic splines with 4 degrees of freedom), sex, and smoking (never or ever).

### Development of a prediction model incorporating mitochondrial heteroplasmy

After determining that mitochondrial heteroplasmy is a predictor for MN, we evaluated whether information on heteroplasmy can improve the prediction of MN using an existing prediction model for MN in individuals with CHIP, the Clonal Hematopoiesis Risk Score (CHRS)^11^. To build upon the CHRS model for CHIP-positive individuals, we first generated the CHRS and CHRS categories by assigning the scores for 8 components (age ≥65 years, presence of cytopenia, red cell distribution width (RDW) ≥15, mean corpuscular volume (MCV) ≥100, presence of high-risk mutation, single *DNMT3A* mutation, number of mutations, and VAF ≥20%) from the original CHRS ^11^. We then added various metrics for heteroplasmy (heteroplasmy count, presence of heteroplasmy, mMSS, heteroplasmy count and mMSS, and presence of heteroplasmy and mMSS) separately to a Cox proportional hazards model with the 8 components of CHRS, sex, smoking (never or ever), and a history of cancer, and tested for model fit using log-likelihood and Akaike information criterion (AIC). We selected the most parsimonious model with the best model fit, which included the presence of heteroplasmy (Y/N) and mMSS as a continuous variable. We developed a score, CHRS-M, by assigning scores for the presence of heteroplasmy and mMSS by rounding the corresponding coefficients from the final model to the nearest 0.5 and adding 1. The coefficient for presence of heteroplasmy was 0.32 and individuals with any heteroplasmy were assigned a score of 1.5. To apply the scoring algorithm to a linear association of mMSS with MN, we estimated the levels of mMSS that correspond to log(HR) of 0.25, 0.75, and 1.25, so that bins of mMSS will be assigned a score of 1, 1.5, 2, and 2.5, respectively (Supplementary Figure 12). The estimated mMSS values were 0.51, 1.52, and 2.54, and thus were divided into 4 categories: <0.51, 0.51–1.52, 1.52–2.54, ≥2.54. However, there was only 1 individual in the ≥2.54 category and, therefore, was collapsed into 3 groups (<0.51, 0.51–1.52, ≥1.52) for final scores. We defined low-, intermediate-, and high-risk categories of CHRS-M by adding 2 to the cut-offs of CHRS; low (≤11.5), intermediate (12–14), and high (≥14.5). We evaluated the 10-year cumulative incidence of MN by CHRS-M category using Fine-Gray method to account for competing risk due to death ^50^. We also performed Cox proportional hazards models to compare the risk of MN between CHRS-M categories. We evaluated the change in sensitivity and specificity compared to CHRS by using net reclassification index (NRI; R package *nricens*) (https://cran.r-project.org/web/packages/nricens/index.html).

In individuals without CHIP, we estimated the HRs for the association between parameters of heteroplasmy and MN using a Cox proportional hazards model adjusting for age (restricted cubic splines with 4 degrees of freedom), sex, smoking (never or ever), and a history of cancer. We first included heteroplasmy count and mMSS in separate models, and to evaluate whether they are independently associated with MN, we then included heteroplasmy count and mMSS in the same model. Finally, we additionally adjusted for other potential biomarkers of MN, including RDW, MCV, and presence of cytopenia.

## Data Availability

UK Biobank data is available through application to the UK Biobank (Application Number 17731).

ARIC WGS is available through dbGap accesstion number phs001211. Due to restrictions based on privacy regulations and informed consent of the participants, data cannot be made freely available in a public repository. Research data requests can be submitted to steering committee, which will be promptly reviewed for confidentiality or intellectual property restrictions and will not unreasonably be refused.

## Code Availability

Code for data cleaning and analysis is available on our github repository: https://github.com/ArkingLab. Documentation on MitoHPC pipeline for DNA Nexus server is available in https://github.com/ArkingLab/MitoHPC/blob/main/docs/DNAnexus_CLOUD.md. Documentation on extracting Mitochondrial and NUMT reads from Google Cloud is available in https://github.com/ArkingLab/MitoHPC/blob/main/docs/GOOGLE_CLOUD.md.

## Acknowledgements

This research was conducted using the UK Biobank Resource under Application Number 17731. This work was supported by National Heart, Lung and Blood Institute, National Institutes of Health (NIH) grants R01HL144569 (D.E.A) and NHLBI: R01HL156144 (L.P.G.). The content is solely the responsibility of the authors and does not necessarily represent the official views of the NIH.

## ARIC Funding

The Atherosclerosis Risk in Communities study has been funded in whole or in part with Federal funds from the National Heart, Lung, and Blood Institute, National Institutes of Health, Department of Health and Human Services, under Contract nos. (75N92022D00001, 75N92022D00002, 75N92022D00003, 75N92022D00004, 75N92022D00005). Studies on cancer in ARIC are also supported by the National Cancer Institute (U01 CA164975, P01CA265748). Building on GWAS for NHLBI-diseases: the U.S. CHARGE consortium was provided by the NIH through the American Recovery and Reinvestment Act of 2009 (ARRA) (5RC2HL102419). Sequencing was carried out at the Baylor College of Medicine Human Genome Sequencing Center (U54 HG003273 and R01HL086694). The content of this work is solely the responsibility of the authors and does not necessarily represent the official views of the National Institutes of Health.

## ARIC Acknowledgments

The authors thank the staff and participants of the ARIC study for their important contributions. Cancer data was provided by the Maryland Cancer Registry, Center for Cancer Prevention and Control, Maryland Department of Health, with funding from the State of Maryland and the Maryland Cigarette Restitution Fund. The collection and availability of cancer registry data are also supported by the Cooperative Agreement NU58DP006333, funded by the Centers for Disease Control and Prevention. Its contents are solely the responsibility of the authors and do not necessarily represent the official views of the Centers for Disease Control and Prevention or the Department of Health and Human Services.

## TOPMed Acknowledgement

Molecular data for the Trans-Omics in Precision Medicine (TOPMed) program was supported by the National Heart, Lung and Blood Institute (NHLBI). Whole genome sequencing (WGS) for the Trans-Omics in Precision Medicine (TOPMed) program was supported by the National Heart, Lung and Blood Institute (NHLBI). WGS for “NHLBI TOPMed: Atherosclerosis Risk in Communities (ARIC) (phs001211) was performed at the Baylor College of Medicine Human Genome Sequencing Center (HHSN268201500015C and 3U54HG003273-12S2) and the Broad Institute for MIT and Harvard (3R01HL092577-06S1). Core support including centralized genomic read mapping and genotype calling, along with variant quality metrics and filtering were provided by the TOPMed Informatics Research Center (3R01HL-117626-02S1; contract HHSN268201800002I). Core support including phenotype harmonization, data management, sample-identity QC, and general program coordination, were provided by the TOPMed Data Coordinating Center (R01HL-120393; U01HL-120393; contract HHSN268201800001I). We gratefully acknowledge the studies and participants who provided biological samples and data for TOPMed. The Genome Sequencing Program (GSP) was funded by the National Human Genome Research Institute (NHGRI), the National Heart, Lung, and Blood Institute (NHLBI), and the National Eye Institute (NEI). The GSP Coordinating Center (U24 HG008956) contributed to cross-program scientific initiatives and provided logistical and general study coordination. The Centers for Common Disease Genomics (CCDG) program was supported by NHGRI and NHLBI, and whole genome sequencing was performed at the Baylor College of Medicine Human Genome Sequencing Center (UM1 HG008898). The Analysis Commons was funded by R01HL131136.

---

**Supplementary Table 1.** Genes included in the CHIP panel.

|  |  |  |  |  |
| --- | --- | --- | --- | --- |
| ASXL1 | EP300 | MPL | SETD2 | ZBTB33 |
| ASXL2 | ETNK1 | NF1 | SETDB1 | ZNF318 |
| BCOR | ETV6 | NPM1 | SF3B1 | ZRSR2 |
| BCORL1 | EZH2 | NRAS | SMC1A |  |
| BRAF | FLT3 | NXF1 | SMC3 |  |
| BRCC3 | GATA2 | PDS5B | SRCAP |  |
| CBL | GNAS | PHF6 | SRSF2 |  |
| CBLB | GNB1 | PHIP | STAG2 |  |
| CEBPA | IDH1 | PPM1D | SUZ12 |  |
| CREBBP | IDH2 | PRPF40B | TET2 |  |
| CSF3R | IKZF1 | PRPF8 | TP53 |  |
| CTCF | JAK2 | PTPN11 | U2AF1 |  |
| CUX1 | KDM6A | RAD21 | U2AF2 |  |
| DNMT3A | KIT | RUNX1 | WT1 |  |
| EED | KRAS | SETBP1 | YLPM1 |  |

**Supplementary Table 2.** Distribution of mMSS in different CHIP subsets.

| Variable |  | UKB | P | ARIC | P |
| --- | --- | --- | --- | --- | --- |
|  |  | Median mMSS<br>(Q1, Q3) |  | Median mMSS<br>(Q1, Q3) |  |
| **CHIP** | **No CHIP**<br>**CHIP** | 0.020 (0.000, 0.157)<br>0.052 (0.006, 0.293) | <0.0001 | 0.033 (0.000, 0.191)<br>0.074 (0.009, 0.502) | <0.0001 |
| **Clone size** | **2% ≤ VAF < 20**<br>**VAF ≥ 20%** | 0.040 (0.005, 0.248)<br>0.128 (0.014, 0.545) | <0.0001 | 0.056 (0.008, 0.436)<br>0.103 (0.010, 0.587) | 0.37 |
| **Number of**<br>**mutations** | **Single**<br>**Multiple** | 0.046 (0.005, 0.275)<br>0.123 (0.013, 0.541) | <0.0001 | 0.067 (0.007, 0.506)<br>0.162 (0.012, 0.444) | 0.60 |
| **Spliceosome**<br>**mutations** | **Other CHIP**<br>**Spliceosome** | 0.048 (0.005, 0.282)<br>0.228 (0.016, 0.689) | <0.0001 | 0.080 (0.010, 0.471)<br>0.039 (0.005, 0.910) | 0.98 |

Abbreviations: ARIC, Atherosclerosis Risk in Communities study; CHIP, clonal hematopoiesis of indeterminate potential; mMSS, mitochondrial local constraint (MLC) score sum; Q1, 25<sup>th</sup> percentile; Q3, 75<sup>th</sup> percentile; UKB, UK Biobank; and VAF, variant allele fraction.

**Supplementary Table 3.** Sensitivity analyses for the associations of heteroplasmy, CHIP, and MN.

|  | Restricted to self-reported White individuals, HR (95% CI) | Restricted to self-reported non-White individuals HR (95% CI) | Restricted to genetically unrelated individuals, HR (95% CI) | Restricted to never smokers, HR (95% CI) | Restricted to individuals without prevalent cancer, HR (95% CI) |
| --- | --- | --- | --- | --- | --- |
| **Number of participants** | 410,313 | 23,991 | 362,132 | 238,086 | 403,732 |
| **Heteroplasmy vs. CHIP** |  |  |  |  |  |
| **None** | Reference | Reference | Reference | Reference | Reference |
| **Heteroplasmy only** | 1.7 (1.5, 2.0) | 1.7 (0.8, 3.7) | 1.8 (1.5, 2.1) | 1.6 (1.3, 2.0) | 1.7 (1.5, 2.0) |
| **CHIP only** | 5.4 (5.6, 6.5) | 2.4 (0.7, 8.3) | 5.7 (4.7, 6.8) | 4.9 (3.8, 6.3) | 5.6 (4.7, 6.6) |
| **Both heteroplasmy and CHIP** | 12.3 (1.1, 14.4) | 6.4 (2.1, 19.3) | 12.6 (10.6, 14.9) | 13.0 (10.3, 16.4) | 12.6 (10.6, 14.8) |
| **Heteroplasmy metrics** |  |  |  |  |  |
| **Heteroplasmy count** | 1.3 (1.2, 1.4) | 1.4 (0.9, 2.4) | 1.3 (1.2, 1.5) | 1.3 (1.1, 1.5) | 1.2 (1.1, 1.4) |
| **mMSS** | 2.6 (2.1, 3.2) | 4.9 (1.5, 16.5) | 2.7 (2.1, 3.3) | 2.9 (2.1, 4.0) | 2.9 (2.3, 3.6) |
| **CHRS category\*\*** |  |  |  |  |  |
| **Number of participants** | 32,066 | 1,531 | 27,998 | 17,122 | 30,654 |
| **Low** | Reference | Reference | Reference | Reference | Reference |
| **Intermediate** | 9.9 (8.0, 12.3) | 7.7 (1.7, 34.6) | 10.5 (8.3, 13.2) | 10.5 (7.7, 14.3) | 10.4 (8.4, 13.0) |
| **High** | 81.7 (62.4, 107.0) | 173.4 (20.5, 1470) | 87.0 (65.2, 115.9) | 106.7 (70.6, 161.3) | 90.6 (68.4, 120.0) |

\*Heteroplasmy count and mMSS were included in the same model. \*\*Analyses restricted to individuals who have CHIP.
All models are adjusted for age (restricted cubic splines with 4 degrees of freedom), sex, smoking (ever or never), and a history of cancer. Because age is a component of CHRS, age was not further adjusted for in the association between CHRS and MN.
Abbreviations: CHIP, clonal hematopoiesis of indeterminate potential; CHRS, clonal hematopoiesis risk score; MN, myeloid neoplasms; and mMSS, mitochondrial local constraint (MLC) score sum.

**Supplementary Table 4.** Participant characteristics stratified by CHRS category in the UK Biobank.

|  | Low | Intermediate | High | *P* |
| --- | --- | --- | --- | --- |
| **Number** | 30,542 | 2,821 | 234 |  |
| **Age, years** | 58.6 (7.6) | 61.7 (7.1) | 64.6 (4.7) | <0.001 |
| **Men, %** | 44.0 | 51.7 | 71.4 | <0.001 |
| **Race, %** |  |  |  | <0.001 |
| **White** | 95.6 | 93.5 | 98.3 |  |
| **Black** | 1.2 | 2.0 | 0.9 |  |
| **Asian** | 1.7 | 2.4 | 0.9 |  |
| **Other** | 1.2 | 1.4 | 0 |  |
| **Ever smoker, %** | 62.0 | 65.6 | 75.6 | <0.001 |
| **CHRS components** |  |  |  |  |
| **Age ≥65 years, %** | 25.2 | 49.3 | 63.2 | <0.001 |
| **CCUS, %** | 5.2 | 29.9 | 58.1 | <0.001 |
| **RDW ≥15, %** | 2.6 | 36.9 | 76.5 | <0.001 |
| **MCV ≥100, %** | 1.2 | 13.9 | 37.2 | <0.001 |
| **High-risk mutation, %** | 2.3 | 33.3 | 85.9 | <0.001 |
| **Single DNMT3A, %** | 32.9 | 6.0 | 1.3 | <0.001 |
| **Number of mutations ≥2, %** | 5.3 | 38.6 | 66.7 | <0.001 |
| **VAF ≥20%** | 12.1 | 47.9 | 87.6 | <0.001 |
| **Heteroplasmy count** | 0 (0 – 1) | 0 (0 – 1) | 1 (0 – 2) | <0.001 |
| **mMSS** | 0 (0 – 0.00) | 0 (0 – 0.07) | 0.02 (0 – 0.60) | <0.001 |

Continuous variables are displayed as either mean (SD) or median (25<sup>th</sup> and 75<sup>th</sup> percentiles). Abbreviations: CCUS, clonal cytopenia of undetermined significance; CHRS, clonal hematopoiesis risk score; MCV, mean corpuscular volume; mMSS, mitochondrial local constrain (MLC) score sum; RDW, red cell distribution width; and VAF, variant allele fraction.

**Supplementary Table 5.** ICD-9 and ICD-10 codes used for the definition of MN and its subtypes in the UK Biobank.

|  | **ICD-9** | **ICD-10** |
| --- | --- | --- |
| **Acute myeloid leukemia** | 205.0, 205.2, 205.3, 206.0, 206.2, 206.9, 207.0, 207.2, 208.0, 208.2 | C92.0, C92.3, C92.4, C92.5, C92.6, C93.0, C93.9, C94.0, C94.2, C94.3, C94.4, C95.0 |
| **Chronic myeloproliferative disease / myeloproliferative neoplasms** | 238.4, 238.7 | D45, D47.1, D47.3, D47.4, C94.6 |
| **Myelodysplastic syndrome** |  | D46.X |
| **Chronic and other myelogenous leukemia** | 205.1, 205.8, 205.9, 206.1, 206.8, 207.8, 208.1, 208.8, 208.9 | D47.5, C92.2, C92.7, C92.9, C93.1, C93.2, C93.7, C94.1, C94.7, C95.1, C95.2, C95.7, C95.9 |

**Supplementary Table 6.** ICD-O-3, ICD-9/10 codes used for the definition of MN and its subtypes in ARIC.

|  | **ICD-9** | **ICD-10 (ICD-O-3)** |
| --- | --- | --- |
| **Acute myeloid leukemia** |  | C42.1 (9861, 9866, 9867, 9871, 9872, 9873, 9891, 9895, 9896, 9931) |
| **Myeloproliferative Neoplasm** | 238.4, 238.71 | C42.0 (9962); C42.1 (9950, 9960, 9962) |
| **Mast Cell Neoplasm** | 238.5 | C42.1 (9741) |
| **Myelodysplastic Syndrome** | 238.72, 238.73, 238.75 | C42.1 (9980, 9982, 9983, 9986, 9987, 9989, 9999), D46.9 |
| **Chronic Myeloid Leukemia** |  | C42.1 (9863, 9875) |
| **Myelodysplastic Syndrome Myeloproliferative Neoplasm Overlap** |  | C42.1 (9945, 9975) |

## Supplementary figure legends

**Supplementary Figure 1.** Representation of the first 15 most frequently mutated genes in (**A**) UKB and (**B**) ARIC.

**Supplementary Figure 2.** Exploratory plot showing the association between different CHIP mutations, heteroplasmy prevalence, and median mMSS. In the UKB (**A**), the mutated genes are limited to those present in more than 10 individuals. In ARIC (**B**), the mutated genes are limited to those present in (**A**), present in more than three individuals, and present in at least one individual with the selected gene and exhibiting heteroplasmy.

**Supplementary Figure 3.** The prevalence of mtDNA complex/region mutations within individuals with heteroplasmy across different CHIP subsets: **A)** CHIP vs No CHIP. **B)** VAF≥20% vs 2%≤VAF<20%. **C)** Multiple mutations vs Single mutation. **D)** Spliceosome mutations vs Other CHIP. The denominator was represented by the total individuals with heteroplasmy from the respective CHIP subset. The mMSS by complex/region among individuals with heteroplasmy across different CHIP subsets: **E)** CHIP vs No CHIP. **F)** VAF≥20% vs 2%≤VAF<20%. **G)** Multiple mutations vs Single mutation. **H)** Spliceosome mutations vs Other CHIP.

**Supplementary Figure 4.** Hazard ratios (95% confidence intervals) for the association between heteroplasmy and incident MN. The adjusted hazard ratios for heteroplasmy count (as categorical and continuous variables) and mMSS in all participants in the UKB (**A**) and in participants with heteroplasmy (**B**). Hazard ratios (95% confidence intervals) were estimated using Cox proportional hazards models adjusting for age modeled as a restricted cubic spline, sex, smoking status, and the presence of prevalent cancer. Abbreviations: CI, confidence interval; HR, hazard ratio; and mMSS, modified mitochondrial local constraint (MLC) sum.

**Supplementary Figure 5.** Hazard ratios (95% confidence intervals) for the association between heteroplasmy and incident MN by mtDNA complex / region. The adjusted hazard ratios for heteroplasmy count (**A**) and mMSS (**B**) by complex / region in the UKB. Hazard ratios (95% confidence intervals) were estimated using Cox proportional hazards models adjusting for age modeled as a restricted cubic spline, sex, smoking status, and the presence of prevalent cancer. Abbreviations: CI, confidence interval; HR, hazard ratio; and mMSS, modified mitochondrial local constraint (MLC) sum.

**Supplementary Figure 6.** Hazard ratios (95% confidence intervals) for the associations of CHRS and CHRS-M categories with subtypes of MN. The analysis was restricted to UKB participants with CHIP. Hazard ratios (95% confidence intervals) were estimated using Cox proportional hazards models with the low-risk group as the reference category. All models are adjusted for age modeled as a restricted cubic spline, sex, smoking status, and the presence of prevalent cancer. Abbreviations: AML, acute myeloid leukemia; CI, confidence interval; CHRS, clonal hematopoiesis risk score; CHRS-M, clonal hematopoiesis risk score with mitochondrial heteroplasmy; HR, hazard ratio; MDS, Myelodysplastic syndrome; MN, myeloid neoplasms; and MPN, myeloproliferative neoplasms.

**Supplementary Figure 7.** Distribution of heteroplasmy count and mMSS by CHRS category in the UKB. (**A**) Proportions of heteroplasmy count (0, 1, 2, 3, and 4+) within each CHRS category. (**B**) Boxplots and scatterplots of the distribution of mMSS by CHRS category. The vertical lines within the box indicate the 25^th^ (Q1), 50^th^, and 75^th^ (Q3) percentiles of the distribution. The horizontal lines indicate the Q1 – 1.5 x (Q3 – Q1) and Q1 + 1.5 x (Q3 – Q1). Values outside the range of the horizontal lines (outliers) are displayed as dots. Abbreviations: CHRS, clonal hematopoiesis risk score; and mMSS, modified mitochondrial local constraint (MLC) score sum.

**Supplementary Figure 8.** Distribution and components of CHRS-M in UKB. (**A**) Histogram of the distribution of CHRS-M. The number of participants corresponding to each CHRS-M score is displayed above the bars. (**B**) Hazard ratios (95% confidence intervals) for each component of CHRS-M and incident MN estimated from a multivariable Cox proportional hazards model including all components of CHRS-M, sex, smoking status, and the presence of prevalent cancer. Abbreviations: CI, confidence interval; CCUS, clonal cytopenia of undetermined significance; CHRS-M, clonal hematopoiesis risk score with mitochondrial heteroplasmy; HR, hazard ratio; MCV, mean corpuscular volume; mMSS, mitochondrial local constrain (MLC) score sum; and RDW, red cell distribution width; and VAF, variant allele fraction.

**Supplementary Figure 9.** Flowchart of UKB participants.

**Supplementary Figure 10.** Flowchart of ARIC participants.

**Supplementary Figure 11.** Comparison of hazard ratios (95% confidence intervals) for the associations of heteroplasmy, CHIP, and incident MN by different VAF cut-offs for heteroplasmy. Hazard ratios (95% confidence intervals) were estimated using Cox proportional hazards models adjusting for age modeled as a restricted cubic spline, sex, smoking status, and the presence of prevalent cancer in UKB. (**A**) Hazard ratios of incident MN by presence of heteroplasmy only, CHIP only, or both heteroplasmy and CHIP, compared to absence of heteroplasmy and CHIP. (**B**) Hazard ratios of incident MN by CHRS category compared to participants without CHIP. (**C**) Hazard ratios of incident MN for heteroplasmy count and mMSS in participants with CHIP. (**D**) Hazard ratios of incident MN for heteroplasmy count and mMSS in participants without CHIP. For (**C**) and (**D**), heteroplasmy count and mMSS were included in the same model. Abbreviations: CHIP, clonal hematopoiesis of indeterminant potential; CI, confidence interval; HR, hazard ratio; mMSS, mitochondrial local constraint (MLC) score sum; and VAF, variant allele fraction.

**Supplementary Figure 12.** Linear association between mMSS and incident MN used to generate CHRS-M. Purple line and band indicate the log(hazard ratio) and 95% confidence interval from a Cox proportional hazards model for incident MN, adjusting for the presence of heteroplasmy, age modeled as a restricted cubic spline, sex, smoking status and the presence of prevalent cancer. The x-axis indicates mMSS, and the y-axis indicates the log(hazard ratio). Blue horizontal lines indicate log(hazard ratio) = 0.25, 0.75, and 1.25. Red vertical lines indicate mMSS = 0.51, 1.52, and 2.54 which correspond to log(hazard ratio) = 0.25, 0.75, and 1.25, respectively. Abbreviations: log(HR), natural log of hazard ratio; mMSS, modified mitochondrial local constraint (MLC) score sum; and MN, myeloid neoplasm.

