## Supplementary Figure 4 for "Mitochondrial heteroplasmy improves risk prediction for myeloid neoplasms"

**A**

| Variable | Categories | Total | Events | Adjusted HR (95% CI) |
| --- | --- | --- | --- | --- |
| Heteroplasmy count | | | | |
| | 0 | 311335 | 633 | Reference |
| | 1 | 99327 | 371 | 1.74 (1.53 to 1.98) |
| | 2 | 19671 | 127 | 2.80 (2.32 to 3.39) |
| | 3 | 3307 | 45 | 5.55 (4.10 to 7.51) |
| | 4+ | 664 | 15 | 8.75 (5.24 to 14.61) |
| Heteroplasmy count | | | | |
| | Continuous | 434304 | 1191 | 1.71 (1.60 to 1.82) |
| mMSS | | | | |
| | Continuous | 434304 | 1191 | 5.55 (4.80 to 6.43) |

**B**

| Variable | Categories | Total | Events | Adjusted HR (95% CI) |
| --- | --- | --- | --- | --- |
| Heteroplasmy count | | | | |
| | 1 | 99327 | 371 | Reference |
| | 2 | 19671 | 127 | 1.61 (1.32 to 1.97) |
| | 3 | 3307 | 45 | 3.19 (2.34 to 4.35) |
| | 4+ | 664 | 15 | 5.03 (3.00 to 8.43) |
| Heteroplasmy count | | | | |
| | Continuous | 122969 | 558 | 1.68 (1.51 to 1.87) |
| mMSS | | | | |
| | Continuous | 122969 | 558 | 4.39 (3.68 to 5.24) |
